# Natural history and impact of *Giardia lamblia* on child growth attainment and associated pathway-specific biomarkers in a Nicaraguan birth cohort

**DOI:** 10.1101/2025.11.09.25339872

**Authors:** Lester Gutiérrez, Nadja A. Vielot, Yaoska Reyes, Roberto Herrera, Christian Toval-Ruiz, Javier Mora, Michael B. Arndt, Rebecca Barney, Robert KM Choy, Filemón Bucardo, Samuel Vilchez, Sylvia Becker-Dreps, Luther A. Bartelt

**Affiliations:** Centro de Investigación de Enfermedades Tropicales (CIET). Facultad de Microbiología. Universidad de Costa Rica, San José, Costa Rica; Department of Family Medicine, University of North Carolina at Chapel Hill, Chapel Hill, NC, USA; Department of Environmental Sciences and Engineering, University of North Carolina at Chapel Hill, Chapel Hill, NC, USA; Universidad Tecnológica La Salle, León, Nicaragua; Laboratory of Helminthology, Faculty of Microbiology, University of Costa Rica, San José, Costa Rica; Atria Health and Research Institute, Seattle, WA, USA; PATH, Seattle, WA, USA; Department of Microbiology and Immunology, University of North Carolina at Chapel Hill, Chapel Hill, NC, USA; Department of Epidemiology, University of North Carolina at Chapel Hill, Chapel Hill, NC, USA; Center for Gastrointestinal Biology and Disease and the Departments of Medicine, University of North Carolina at Chapel Hill, Chapel Hill, NC, USA; Division of Infectious Diseases, Department of Medicine, University of North Carolina at Chapel Hill, Chapel Hill, NC, USA

## Abstract

**Background:** *Giardia lamblia* (*Giardia*) is one of the most common intestinal parasitic infections globally, with an estimated 280 million symptomatic infections annually. In children from low-and middle-income countries (LMICs), *Giardia* is highly prevalent and has been associated with loss of intestinal barrier function, nutrient-metabolic dysregulation, and linear growth impairment, but specific mechanisms linking *Giardia* to these outcomes remain poorly understood.

**Methods and results:** We used data and samples from a subset of 76 children in a longitudinal birth cohort in Nicaragua to evaluate the natural history and geospatial distribution of *Giardia* infections, child growth outcomes (weight-for-age [WAZ] and length-for-age [LAZ] z scores), and relationships with established biomarkers of inflammation, intestinal damage, and growth-signaling. During the first 36 months of life, we tested 2,305 stools (1,903 surveillance stools and 402 diarrheal stools) for *Giardia* by qPCR. The incidence of *Giardia*-positive stools was 59.6 per 100 child-years. Any detection of *Giardia* was associated with a reduction in LAZ at 36 months of life (β:-0.16, *P*=0.042). This effect increased when considering persistent or recurrent *Giardia* detections (β:-0.26, *P*=<0.001) as well as living in a high-density *Giardia* detection area (β:-0.44, P=<0.001). Among intestinal markers, *Giardia* was only associated with lower median fecal neopterin (a marker of chronic intestinal T cell activation) at 24 and 36 months of age. Among serum systemic biomarkers measured at 24 months, *Giardia* detections were associated with indicators of intestinal epithelial cell damage (higher median Intestinal Fatty Acid Binding Protein (*P*=0.002) and Anti-FliC IgA (P=0.033), and reduced growth-signaling hormone (lower median Insulin-like Growth Factor (IGF-1) (*P*=0.005).

**Conclusion:** *Giardia* detection was negatively associated with linear growth in an exposure-dependent manner. Simultaneously, *Giardia* associates with diminished serum growth-signaling hormones. Patterns of serum and fecal intestinal biomarkers suggest that *Giardia-*mediated epithelial disruption is dissociated from markers of intestinal inflammation.

**Author Summary:** Although *Giardia* was recognized as a pathogen by the World Health Organization in 1981, its long-term effects on child health, particularly in endemic areas, remain poorly understood. One major challenge is that most *Giardia* infections are asymptomatic and go unreported, leading to an underestimation of not only its true burden but also its association with child health outcomes. While a link between *Giardia* and impaired linear growth is becoming clearer in large prospective child cohorts, considerable variability persists in the strength and consistency of this association across studies. Moreover, the mechanistic pathways underlying long-term sequelae in children remain elusive. To contribute to this field, we leveraged a birth cohort with longitudinal biosampling to study *Giardia* infections and their impact on child growth. We found that *Giardia* infection was negatively associated with length-for-age and Insulin-like Growth Factor 1 (IGF-1) levels, positively associated with biomarkers of intestinal epithelial disruption, yet uncoupled from markers of systemic inflammation and intestinal inflammation. The association with poor linear growth, epithelial disruption, and reduced growth signaling from intestinal inflammation which is a unique characteristic of *Giardia*, requires further investigation of its pathophysiology.

## Introduction

*Giardia lamblia* (also known as *Giardia duodenalis* or *Giardia intestinalis*) is globally distributed intestinal protozoan parasite, primarily transmitted through contaminated water. An estimated 280 million episodes of symptomatic *Giardia* occur globally each year [1], and the majority of which are concentrated in low-and middle-income countries (LMICs). An even greater number of asymptomatic infections occur, yet are infrequently reported. *Giardia* can result in a broad spectrum of clinical outcomes, ranging from asymptomatic carriage to symptomatic diarrhea, conventionally characterized by abdominal cramping, bloating, steatorrhea, and weight loss. Chronic gastrointestinal disturbances also occur [2,3].

In LMICs where *Giardia* is highly endemic, its detection in the stool (regardless of symptoms) has been associated with increased intestinal permeability and reduced anthropometric measurements such as length-for-age [LAZ] and to a lesser extent weight-for-age [WAZ] in colonized children [4]. Notably, the Malnutrition and Enteric Disease study (MAL-ED) identified *Giardia* as one of the top five contributors to linear growth faltering in children under two years of age [5]. Although longitudinal studies have increasingly supported an association between *Giardia* infection and impaired linear growth [5–10], the magnitude of its impact on child growth remains inconsistent across different geographic regions and populations. This variability persists even in multi-site studies following an identical protocol, suggesting that study design alone does not account for these differences [11]. Further research is needed to investigate host and environmental factors, as well as the role of persistent and recurrent *Giardia* infections, to better understand their contribution to individual-level variations in child growth and development.

The pathogenesis underlying growth failure following *Giardia* infection remains poorly understood. Different potential mechanistic pathways have been described [12]. Evidence suggests that linear growth faltering and intestinal barrier dysfunction may be independent of gut inflammation, as measured by fecal biomarkers commonly associated with Environmental Enteric Dysfunction (EED), such as neopterin and myeloperoxidase [11]. Multiple studies have reported an absence of conventional intestinal markers of inflammation in children with concurrent *Giardia* detection. [4,13,14], yet simultaneously findings of increased intestinal permeability and disruption of nutrient-metabolic pathways [4,11]. Experimental models in gnotobiotic mice have recently demonstrated *Giardia-*mediated impaired growth and inadequate nutrient absorption in the absence of prototypic EED-like chronic lymphocytic inflammation [12]. Further research is required to elucidate the precise mechanisms by which *Giardia* contributes to the growth impairment.

We followed a cohort of 76 Nicaraguan children from birth through 36 months of age and captured high-resolution weekly longitudinal growth data, monthly surveillance stool sampling, episodic diarrhea samples, and 49 serum samples to determine the contributions of *Giardia* on linear growth trajectories. Additionally, we profiled indicators of systemic and intestinal inflammation and gut epithelial damage, by testing for associations with asymptomatic infections, and for dose-dependence (eg. persistent or recurrent *Giardia* infections) with those outcomes.

## Methods

### Ethics statement

The study was approved by the Ethical Committee for Biomedical Research (CEIB) “Dr. Uriel Guevara Guerrero” of the National Autonomous University of Nicaragua (UNAN) at León (Acta number 2–2017; FWA00004523 / IRB00003342) and the Institutional Review Board of the University of North Carolina at Chapel Hill (study number 16– 2079). Written informed consent was obtained from a parent or legal guardian of each participant prior to enrollment in the study.

In addition, consent for sample storage and future unspecified analyses was obtained from the caregivers on behalf of their children.

### Participants

This study included a subset of children enrolled in the Sapovirus-Associated GastroEnteritis (SAGE) birth cohort study between June 12, 2017 and July 31, 2018 in León, Nicaragua [15]. In the SAGE cohort, 443 children were visited weekly in their households from birth until 36 months of age to surveil for acute gastroenteritis (AGE) episodes (defined as diarrhea and/or vomiting) and collect epidemiological data as we described previously [15,16]. Each month, mothers were asked to provide data regarding the birth characteristics like sex, mode of delivery, mean birth length and weight, socioeconomic indicators, and breastfeeding data. Socioeconomic index was created following Peña *et al.* criteria for this setting [17]. Participant height and weight were measured at monthly intervals along the cohort (Fig. S1). Global Positioning System (GPS) coordinates were also recorded and used for spatial *Giardia* density analysis.

Available resources for *Giardia* detection and bioassays restricted the subset of children in this study to 76 participants. We therefore first restricted the sample to size to only those children who contributed ≥ 90% of expected surveillance samples (≥27 stools over 36 months of follow-up) (N=107). Among these 107 children, we randomly selected 76. Baseline demographic and epidemiological characteristics of these 76 children did not differ significantly from those of the remaining cohort (Table S1).

### Specimen collection

Routine stool samples for each child were collected monthly from birth to 24 months of age, and also at 27, 30, 33, and 36 months of age (Fig. S1). Additionally, stool samples were collected from each of the reported AGE episodes, which were defined as at least three loose stools in a 24-hour period, a change in the consistency of stools (bloody, very loose, or watery), or the presence of vomiting, following at least three symptom-free days [15]. When AGE stools were collected at the same time corresponding to the routine sampling, these samples were classified as AGE and excluded from routine stool category. Both routine and AGE stool samples were stored at-20°C raw and also in aliquots of 1:10 (100 mg/ml) stool suspension in phosphate-buffered saline (PBS) at-20°C until processing. Blood collections were performed by venipuncture at 24 months, and serum was extracted by centrifugation and stored at-80°C until processing.

### Molecular detection of *Giardia*

To identify *Giardia* using Real-Time PCR, 200μl of the 1/10 stool suspension was used to extract DNA using the QIAamp Fast DNA Stool Mini Kit (Cat No./ID: 51604) and by following the manufacturer’s instructions. Stool suspension was initially treated with acid-washed glass beads (0.5 mm; Sigma) and vortexed for 2–5 min, as described by Stroup SE *et al* [18], to increase cyst lysis and DNA extraction. Real Time-PCR was performed to identify *Giardia* in AGE stool samples using the protocol described by Verweij JJ *et al* that target the small subunit ribosomal (SSU) gene (18S-like) from *Giardia* (GenBank accession no. M54878) [19]. In brief, 0.2M of the forward and reverse primers and probe (*Giardia*-80F 5’-GAC GGC TCA GGA CAA CGG TT-3’, *Giardia*-127R 5’-TTG CCA GCG GTG TCC G-3’, *Giardia*-105T FAM-5’-CCC GCG GCG GTC CCT GCT AG-3’) were added to a PCR reaction mix consisting of 3 μL of DNA, 12.5 μL of Bio-Rad iQ Multiplex Powermix (Bio-Rad Laboratories, Hercules, CA, USA) and nuclease-free water to a final volume of 25μl. PCR conditions were 95°C 10 min and 45 cycles: 95°C 10 s, 60°C 1 min (signal reading). Real-time PCR was performed using the Bio-Rad CFX96 Touch Real-Time PCR Detection System. The real-time PCR was considered positive if the cycle threshold (Ct) was of ≤35. Only one sample resulted with a Ct value between 36-45. That child otherwise had recurrent *Giardia* positive results. There was therefore neglibible risk for misclassification bias based on the Ct threshold cut-off. Carryover contamination was controlled by using nuclease-free water, during DNA purification and real-time PCR. The positive control was ATCC Quantitative Synthetic DNA from *Giardia lamblia* PRA-3006SD with a Ct of 30±2.

### Biomarkers for pathways of growth-faltering

#### Intestinal Biomarkers

ELISA for fecal Neopterin (NEO), Myeloperoxidase (MPO), and Regenerating family member 1β (Reg-1β) were performed at 12, 24, and 36 months per child (Fig S1). ELISA kits were purchased from commercial vendors (NEO: GenWay Biotech Inc, San Diego, CA. MPO: Immunodiagnostik AG, Stubenwald-Allee, Bensheim, Germany. Reg-1β: RayBiotech Inc, Norcross, GA) and run per the manufacturer’s directions. Stool samples were diluted 1:100 with assay buffer before use in the NEO ELISA kit, 1:500 for use in the MPO ELISA kit, and 1:1000 for use in the Reg-1β ELISA kit. Final concentrations were obtained using the standard curve method. Data were analyzed both raw data and normalized to fecal total protein measured by Pierce BCA Protein Assay Kits performed according to the manufacturer’s directions.

#### Systemic Biomarkers

Serum samples collected at 24 months of age were tested to determine concentrations of intestinal fatty acid binding protein (I-FABP), soluble CD14 (sCD14), insulin-like growth factor 1 (IGF-1), fibroblast growth factor 21 (FGF21), alpha-1 acid glycoprotein (AGP), C-reactive protein (CRP), soluble transferrin receptor (sTfR), retinol binding protein 4 (RBP4) as measured by the MEEDAT 11-plex ELISA (Q-Plex™ Human Environmental Enteric Dysfunction 11-plex, Quansys Biosciences, USA) as described by Arndt *et al* [20] (Table S2). Additionally, serum samples were also analyzed for Anti-Flic IgA using ELISA commercial kit (SunLong Biotech Co. LTD, Hangzhou, China) according to the commercial kit’s instructions.

### Statistical analysis

All statistical analyses were performed using R software (version RStudio 2023.09.0+463; R Foundation for Statistical Computing, Vienna, Austria). First, the birth characteristics like sex, mode of delivery, mean birth length and weight, socioeconomic indicator, and breastfeeding data of the children were described using percentages with standard deviations or medians with interquartile ranges (IQR). Then, we calculated the frequency and incidence rate (events/100 child-years) for symptomatic and asymptomatic *Giardia* infection throughout the cohort, as well as for persistent and recurrent infection in infected children. A persistent infection in this cohort was defined as two or more consecutive stools with a *Giardia* positive qPCR result, and a recurrent infection was defined as two or more non-consecutive stools with a positive *Giardia* qPCR. Next, we performed a Kernel Density Estimation (KDE) analysis using ArcGIS Software v10.5 to assess the density of *Giardia* infections within the study area.

We assessed correlates of *Giardia* detection, persistent infection, and recurrent infection using a Pearson’s chi-square test or Fisher’s exact test for cell sizes <5 for categorical variables. Potential correlates of interest identified in prior literature included age, gender, water source, delivery mode, floor type, and sanitation type and breastfeeding. Categorical characteristics associated with *Giardia* detection below the P = 0.1 type I error level were evaluated for independent associations with *Giardia* detection using a multivariable Cox proportional hazards model to estimate the adjusted relative hazard and 95% confidence interval (CI) for *Giardia* infection.

Potential confounders for exposure-outcome were identified through analysis of directed acyclic graphs (Fig. S2)

Monthly weight and length were used to assess growth velocities for length-for-age (LAZ) and weight-for-age (WAZ) using the WHO growth standards for girls and boys [21]. Effect on the growth velocities was performed at 2 levels. First, the Mann-Whitney U test was used to assess the effect of *Giardia* detection on growth outcomes (LAZ and WAZ) using the endpoint measurements at 24 and 36 months old between any precedent *Giardia-*detected and non-detected children throughout the cohort. Second, we conducted linear regression analyses using generalized estimating equations (GEE) to assess the effect of *Giardia* infection on LAZ and WAZ. As an exporatory analysis, we conducted linear regression to test for associations between systemic and fecal biomarker concentrations with contemporaneous LAZ and WAZ measurements.

Effects were estimated using monthly longitudinal LAZ and WAZ measurements for “any *Giardia*”, “recurrent event”, and “persistent event” (Fig. S3). The model for any *Giardia* infection began at the first positive *Giardia* infection (red circle “b”), using the value one month prior as the baseline (green triangle “a”). The recurrent event models began at the second non-consecutive *Giardia-positive* stool, with the baseline defined as one month prior to this event.

The persistent infection model started at the second consecutive positive stool (red circle “d”), with the baseline also defined as one month prior to this event. All models were adjusted for child’s age at infection, mode of delivery, sex, socioeconomic status, breastfeeding, and episodes of diarrhea during the same period. A *P* value <0.05 was considered statistically significant.

Furthermore, fecal biomarker levels at 24 and 36 months, as well as systemic biomarkers at 24 months of age, were analyzed using the Mann–Whitney U test, comparing children who had been infected with *Giardia* by those ages with those who had not. Finally, Spearman’s correlation was used to evaluate the relationship between the number of *Giardia* infections and changes in biomarkers, as well as their impact on child growth, measured by WAZ and LAZ. The interpretation of Spearman’s rho coefficient followed the criteria proposed by Schober *et al*. [22]: where values between 0.00 and 0.10 were considered negligible; from 0.10 to 0.39 as weak; 0.40 to 0.69 as moderate; from 0.70 to 0.89 as strong, from 0.90 to 1.00 as very strong correlations. A P value <0.05 was considered statistically significant. Images were generated using GraphPad Prism V7 (GraphPad Software Inc).

## Results

### Epidemiological characteristics of population

Of the 76 children included in this study, 31 (41%) were female, 36 (47%) born vaginally, 73 (96%) of children received non-exclusive breastfeeding with a median duration of 16.5 (IQR: 5.7, 31.2) months (Table 1). Exclusive breastfeeding was short-lived (median of 3.1 weeks, IQR: 0.5, 5.7). Twenty-eight (38%) of children lived in houses not having basic needs met (poverty score ≥2) (Table 1), where 21 (28%) had no indoor toilet and 20 (26%) had an earthen floor at home. Only 5 (7%) of houses did not have potable water at home.

**Table 1.** Epidemiological characteristics of children in the sub-cohort study (n=76 children).

| Characteristics | n (%) or median (IQR) |
| --- | --- |
| *Birth characteristic* |  |
| Sex (%Female) | 31 (41) |
| Mode of delivery (%Vaginal) | 36 (47) |
| Mother's age at birth (years) | 20 (23, 26) |
| Mean birth weight (in kg) | 3.2 (3.0, 3.5) |
| Mean birth length (in cm) | 51.3 (50.3, 52.6) |
| *Socioeconomic indicator* |  |
| Poverty index (% poor or extremely poor) (2 missing)<sup>a</sup> | 28 (37) |
| Sanitation type (%Latrine) | 21 (28) |
| Floor-type (%Earthen) | 20 (26) |
| Water resources (%Non-potable at home) | 5 (7) |
| Maternal education attainment (3 missing) |  |
| Completed primary education or less | 20 (27) |
| Completed any secondary education | 53 (73) |
| *Breastfeeding* |  |
| Ever Breastfed | 73 (96) |
| Median duration of breastfeeding (in months) | 16.5 (5.7, 31.2) |

<sup>a</sup> Poverty index was determined according to Peña *et al* [17].

### Incidence of *Giardia* infections

Among the 76 children, 2305 stool samples were collected and tested for *Giardia* by qPCR. 1903 were collected for routine monthly surveillance, and 402 were AGE-stools. A total of 44 of (57.89%) of the 76 children experienced at least one *Giardia* detection. The overall incidence of *Giardia* detections was 59.6 per 100 child-years (95% CI, 49.6–69.7). Incident *Giardia* detection increased from the first year (15.8 [95% CI, 6.8–61.3] per 100 child-years) to the second year (123.7 [95% CI, 98.7–148.7] per 100 child-year) and was 46.1 [95% CI, 30.8–61.3] in the third year (Table 2). Incidence of *Giardia* detection was expectedly higher in routine stools compared with diarrheal-stools (35.5 [95% CI, 27.7–43.2] vs 24.7 [95% CI, 18.2–31.3] per 100 child-years) (Table 2 and Fig. 1). Throughout the follow-up, 18 of the 44 children who were ever infected with *Giardia* (40.9%) had a persistent infection, while 10/44 (22.7%) experienced a reinfection event (Table 2 and Fig. S4).

**Figure 1.**
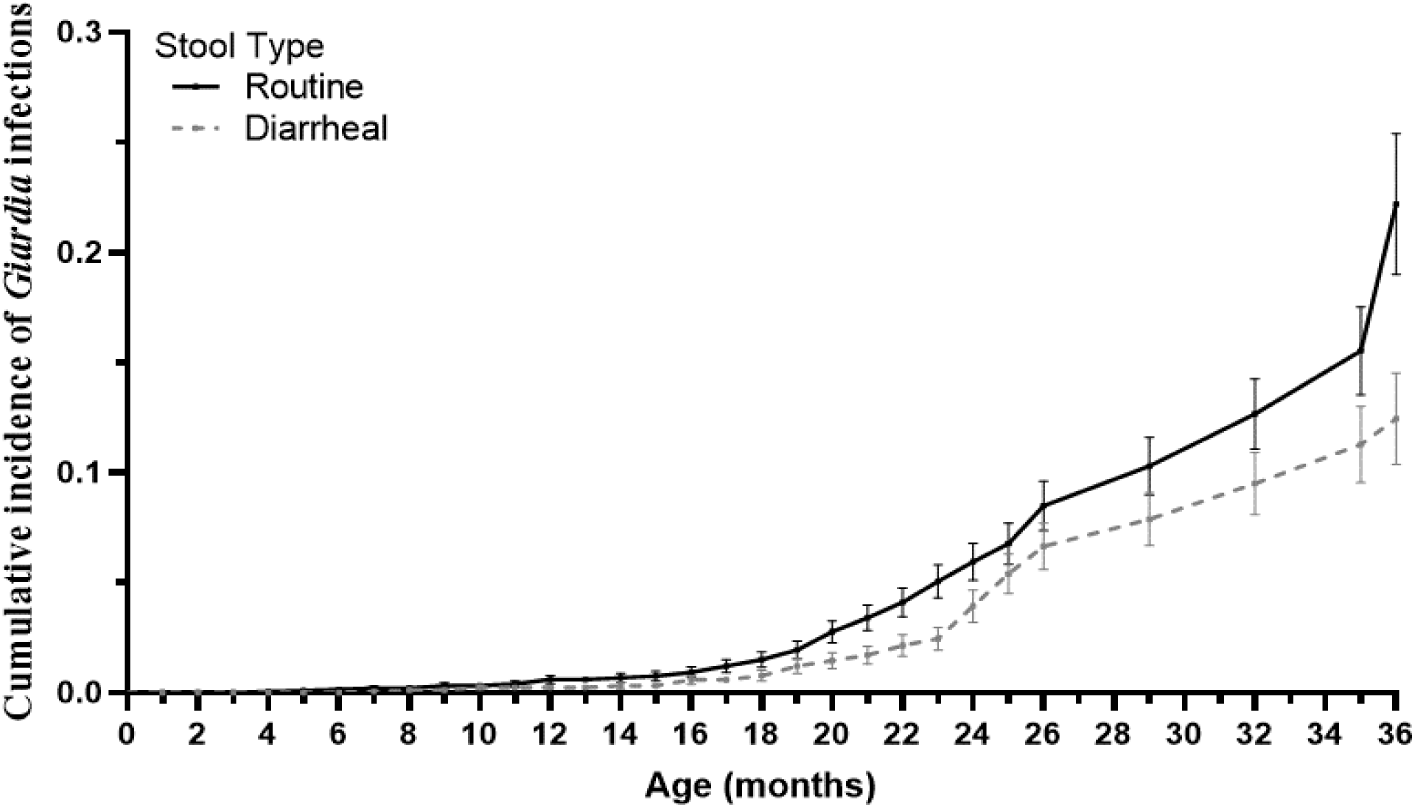
Cumulative incidence of *Giardia* detection in routine (n=1903) and diarrheal stool (n=402) samples among the 76 children.

**Table 2.** Incidence of *Giardia* detection in routine and diarrheal stools in children from León (n=76 children)

| Events | Incidence<sup>a</sup> |
| --- | --- |
| Overall Incidence of *Giardia*-Positive Stool (n=76) | 59.6 (49.6-69.7) |
| Overall Incidence of *Giardia* in the 1<sup>st</sup> year* | 15.8 (6.8-61.3) |
| Overall Incidence of *Giardia* in the 2<sup>nd</sup> year* | 123.7 (98.7-148.7) |
| Overall Incidence of *Giardia* in the 3<sup>rd</sup> year** | 46.1 (30.8-61.3) |
| Incidence of *Giardia* in routine stools (n=76) | 35.5 (27.7-43.2) |
| Incidence of *Giardia* in diarrheal stool (n=74) | 24.7 (18.2-31.3) |
| Incidence of New *Giardia* infections<sup>β</sup> (n=76) | 34.6 (27.0-42.2) |
| Incidence of *Giardia* Reinfection Events (n=44)<sup>1</sup> | 46.3 (17.6-75.1) |
| Incidence of Persistent *Giardia* Infections (n=44)<sup>§</sup> | 40.9 (22.1-59.8) |

<sup>a</sup>Incidence per 100 child-years. \*Incidence in the 1<sup>st</sup> and 2<sup>nd</sup> years was calculated using monthly routine stool samples. \*\* Incidence in the 3<sup>rd</sup> year was calculated using quarterly routine stool samples. <sup>β</sup>New *Giardia* episodes were defined if *Giardia* was detected positive in surveillance or diarrheal stools, and the most recent previous stool tested negative for *Giardia*. <sup>1</sup>*Giardia* re-infection was defined as a child having more than one new *Giardia* episode in non-consecutive stool samples. <sup>§</sup>Persistent *Giardia* infections were defined as two or more consecutive monthly stool samples that tested positive for *Giardia*.

### Characteristics related to *Giardia* infections

Birth, socioeconomic, and nutritional characteristics were compared between children who had at least one *Giardia* detection (n=44) and those with no *Giardia* detection during cohort follow-up (n=32). *Giardia* detections were more frequent among children living in houses not having basic needs met (*P*=0.013), in houses using latrines for sanitation (as compared to having an indoor toilet) (*P*=0.012), and those with an earthen floor (*P*=0.001) (Table 3). The proportion of those socioeconomic indicators was greater among those with >2 detections compared to ≤2 detections (Table 4). Birth characteristics (including sex, mode of delivery, and gestational age), history of an AGE episode of any etiology, and breastfeeding duration were not associated with *Giardia* detection. After adjusting for potential confounders, living without basic needs met (aHR, 2.30 [95% CI, 1.23–4.28]) and having an earthen floor in the home (aHR, 2.90 [95% CI, 1.58–5.50]) remained significantly associated with an increased risk of *Giardia* detection (Table 5).

**Table 3.** Epidemiological characteristics of children with and without *Giardia* infections detected in the first three years of life (n=76 children).

| Characteristics | n (%) or median (IQR) |  | P value* |
| --- | --- | --- | --- |
|  | Children with *Giardia* detection (n=44) | Children without *Giardia* detection (n=32) |  |
| *Birth characteristic* |  |  |  |
| Sex (%Female) | 17 (38.6) | 14 (43.8) | 0.654 |
| Mode of delivery (%Vaginal) | 20 (45.5) | 16 (50.0) | 0.695 |
| Mother's age at birth (years) | 23 (20, 25) | 25 (22, 28) | 0.066 |
| Mean birthweight (in kg) | 3.2 (3.0, 3.5) | 3.1 (2.9, 3.5) | 0.185 |
| *AGE history* |  |  |  |
| Mean of total diarrhea at 3 years | 5 (3, 8) | 4 (3, 7) | 0.722 |
| *Socioeconomic indicator* |  |  |  |
| **Poverty index (% poor or extremely poor) (2 missing)** | **21 (50)** | **7 (21.9)** | **0.013** |
| **Sanitation type (%Latrine)** | **17 (38.6)** | **4 (12.5)** | **0.012** |
| **Floor-type (%Earthen)** | **18 (40.9)** | **2 (6.3)** | **0.001** |
| Water resources (%Non-potable at home) | 4 (9.1) | 1 (3.1) | 0.300 |
| *Maternal education attainment (3 missing)* |  |  |  |
| Completed primary education or less | 11 (26.2) | 9 (29.0) | 0.788 |
| Completed any secondary education | 31 (73.8) | 22 (71.0) | 0.788 |
| *Nutrition* |  |  |  |
| Ever Breastfeeding | 42 (95.5) | 31 (96.9) | 0.754 |
| Breastfeeding durations (in months) | 13 (4, 32) | 21 (8, 31) | 0.415 |
| Median WAZ at birth | 1.22 (0.53, 1.78) | 1.13 (0.39, 1.66) | 0.394 |
| Median LAZ at birth | 0.65 (0.05, 1.55) | 0.71 (0.09, 1.66) | 0.712 |

AGE: acute gastroenteritis. $\alpha$ Poverty index was determined according to Peña *et al* [17]. \*Pearson's chi-square test or Fisher's exact test for cell sizes <5 for categorical variables. Mann-Whitney U test was used for numerical variables

**Table 4.** Epidemiological characteristics of children with less or equal and more than 2 of *Giardia* infections detected during the first three years of life, compared to children without *Giardia* infection.

| Epidemiological characteristics | Children without *Giardia* detection (n=32)* Ref. | Children with $\leq 2$ *Giardia* infections (n=23)* | P. value<sup>1</sup> | Children with $> 2$ *Giardia* infections (n=21)* | P. value<sup>1</sup> |
| --- | --- | --- | --- | --- | --- |
| *Birth characteristic* |  |  |  |  |  |
| Sex (%Female) | 14 (43.8) | 8 (34.8) | 0.503 | 9 (42.9) | 0.949 |
| Mode of delivery (%Vaginal) | 16 (50.0) | 12 (52.2) | 0.874 | 8 (38.1) | 0.394 |
| Mother's age at birth (years) | 25 (22, 28) | 23 (19, 25) | 0.240 | 23 (20, 26) | 0.569 |
| Mean birthweight (in kg) | 3.1 (2.9, 3.5) | 3.3 (3.0, 3.7) | 0.261 | 3.2 (3.0, 3.6) | 0.501 |
| *AGE history* |  |  |  |  |  |
| Median of total diarrhea at 3 years | 4 (3, 7) | 6 (4, 8) | 0.070 | 5 (2, 7) | 0.763 |
| *Socioeconomic indicator* |  |  |  |  |  |
| **Poverty index (% poor) (2 missing)<sup>a</sup>** | 7 (21.9) | 9 (40.9) | 0.132 | **12 (60.0)** | **0.005** |
| **Sanitation type (%Latrine)** | 4 (12.5) | **9 (39.1)** | **0.022** | **8 (38.1)** | **0.029** |
| **Floor-type (%Earthen)** | 2 (6.3) | **8 (34.8)** | **0.007** | **10 (47.6)** | **<0.001** |
| Water resources (%Potable at home) | 31 (96.9) | 19 (82.6) | 0.069 | 21 (100) | 0.413 |
| *Maternal education attainment (3 missing)* |  |  |  |  |  |
| Completed primary education or less | 9 (29.0) | 4 (17.4) | 0.322 | 7 (36.8) | 0.566 |
| Completed any secondary education | 22 (71.0) | 19 (82.6) | 0.322 | 12 (63.2) | 0.566 |
| *Nutrition* |  |  |  |  |  |
| Ever Breastfeeding | 31 (96.9) | 22 (95.7) | 0.811 | 20 (95.2) | 0.760 |
| Breastfeeding duration (in months) | 21 (8, 31) | 8 (4, 28) | 0.109 | 24 (5, 35) | 0.942 |
| Mean WAZ at birth | 1.13 (0.39, 1.67) | 1.51 (0.85, 1.87) | 0.116 | 0.99 (0.19, 1.58) | 0.841 |
| Mean LAZ at birth | 0.71 (0.09, 1.66) | 0.65 (-0.12, 1.51) | 0.804 | 0.65 (0.14, 1.67) | 0.709 |

Ref: Reference group. \*n (%) or median (IQR). <sup>1</sup>Compared to children without *Giardia*. NA: not applicable. AGE: acute gastroenteritis.
<sup>a</sup>Poverty index was determined according to Peña *et al* [17]. <sup>1</sup>Pearson's chi-square test or Fisher's exact test for cell sizes $< 5$ for categorical variables. Mann-Whitney U test was used for numerical variables

**Table 5.** Association between selected characteristics and *Giardia* detection in a birth cohort study (n=76 children)

| Characteristic<sup>§</sup> | Crude |  | Adjusted* |  |
| --- | --- | --- | --- | --- |
|  | HR | 95% CI | HR | 95% CI |
| **Poverty index (% poor) (2 missing)<sup>a</sup>** | **2.31** | **1.26, 4.25** | **2.30** | **1.23, 4.28** |
| Sanitation type (%Latrine) | 2.35 | 1.28, 4.33 | 1.35 | 0.62, 2.95 |
| **Floor-type (%Earthen)** | **2.90** | **1.58, 5.33** | **2.51** | **1.15, 5.50** |

<sup>§</sup>Model adjusted was created using all categorical variables with a $P < 0.1$ in bivariate predictors from Tables 3 and 4.
\*Adjusted analyses were not performed for non-significant variables. Adjusted models included potential confounders for each variable identified through analysis of causal diagrams. Poverty index was adjusted for gestational age, water source, and delivery mode; floor type was adjusted for water source and sanitation type; sanitation type was adjusted for floor type and water source.
HR: Hazard Ratio. CI: Confidence Interval. <sup>a</sup>Poverty index was determined according to Peña *et al* [17].

### Seasonality and Geospatial distribution

*Giardia* was detected across all months of the year (Fig. S5A), with no statistically significant difference between the prevalence of 7.6% in the dry season (November-April) vs 5.23% in the rainy season (May – October) (P=0.101; Fig. S5B). Geospatially, *Giardia* was detected throughout the area studied (Fig. S5C), and the Kernel Density Estimation (KDE) analysis performed demonstrates a single zone of high density of *Giardia* infections, with an estimated value of 22 events per km² (Fig. S5D) in 10 children living inside this area, in which the use of latrine (6/10) and poverty index (5/10) was common. This indicates a marked concentration of events in this specific area, with no other significant density clusters identified across the study region.

### Effects on growth

Growth outcomes were first evaluated using endpoint anthropometric measurements at 24 and 36 months of age in children with at least one *Giardia* detection compared to those without any detection during the cohort. Children with *Giardia* detection showed significantly lower LAZ at 24 (median:-0.93 vs-0.13, P=0.006) and 36 months (median:-1.18 vs-0.50, P=0.007) (Fig. 2A). No significant differences were observed in WAZ at 24 (median:-0.26 vs-0.01, P=0.253) or 36 months of aged (median:-0.31 vs 0.01, P=0.105) (Fig. 2B). Regression models adjusted for age, mode of delivery, sex, socioeconomic status, breastfeeding, and episodes of diarrhea among the 44 *Giardia*-infected children (Table S3) estimated that *Giardia* detection was associated with reduced LAZ at 36 months among children with: any *Giardia* detection (β:-0.16, 95% CI:-0.01,-0.32, P = 0.042) (Fig 3A), any persistence or recurrent infections (β:-0.26, 95% CI:-0.11,-0.41, *P* = <0.001), and more than two *Giardia* detections during the surveillance period (β:-0.29, 95% CI:-0.14,-0.45, *P* = <0.001) (Fig 3A). The geospatial distribution of *Giardia* density was also associated with LAZ. Although reductions in LAZ were observed in children both inside and outside high-density areas, the effect was greater among those living in high-density areas, with a reduction of 0.44 units in LAZ (β:-0.44, 95% CI:-0.18,-0.69, *P* = <0.001), as compared to those living in lower density areas, who had a reduction of-0.19 units (β:-0.19, 95% CI:-0.03,-0.35, *P* = 0.022) (Fig 3A). No differences were observed in WAZ (Fig. 3B).

**Figure 2.**
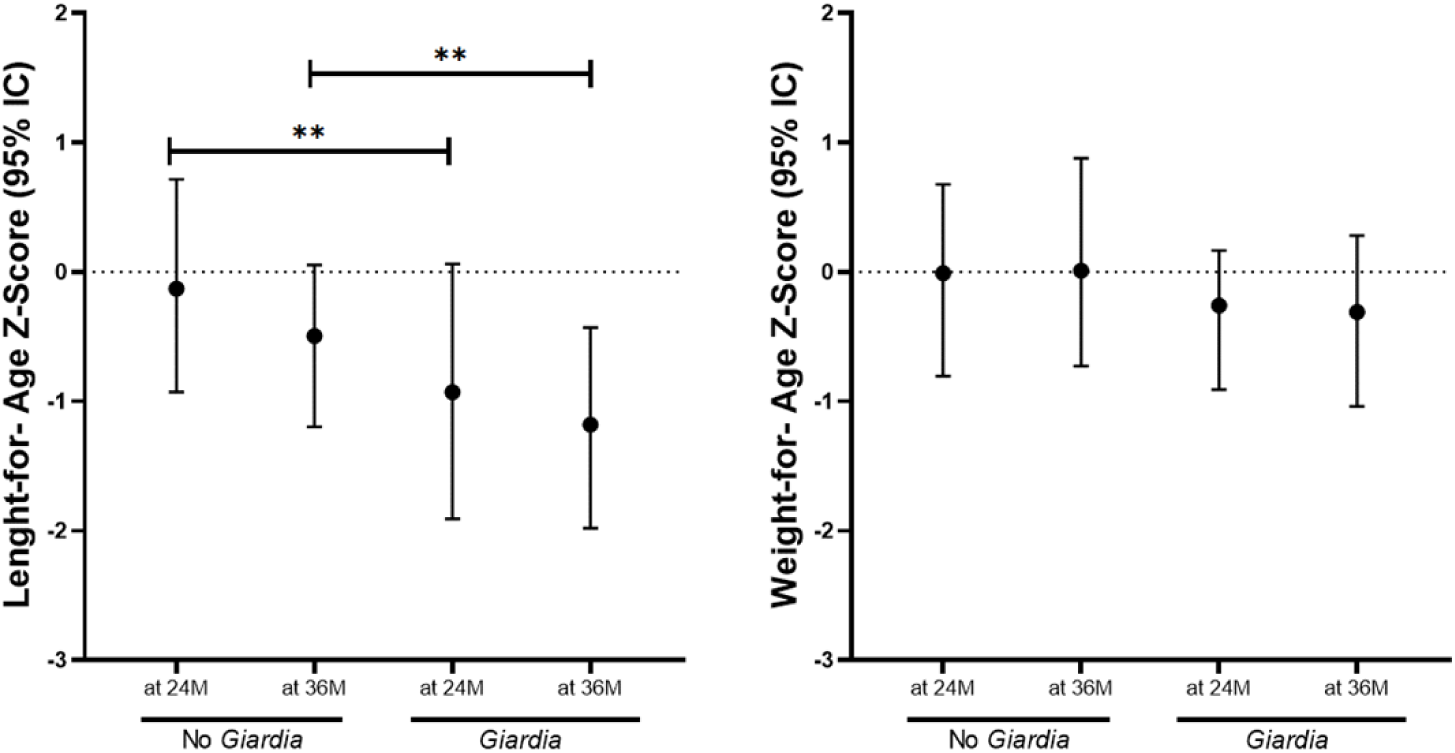
Length-for-age (LAZ) and weight-for-age (WAZ) Z-score at 2 (A) and 3 years old. **(B)** in children experienced at least once *Giardia* detection at 24 (24M) (*Giardia*, n=35) and 36 (36M) months of age (n=44), and children without *Giardia* detection at 24M (No *Giardia*, n=41) and 36M (n=32). \**P<*0.050 to 0.010, \*\**P<*0.010 to 0.001, \*\*\**P<*0.001.

**Figure 3.**
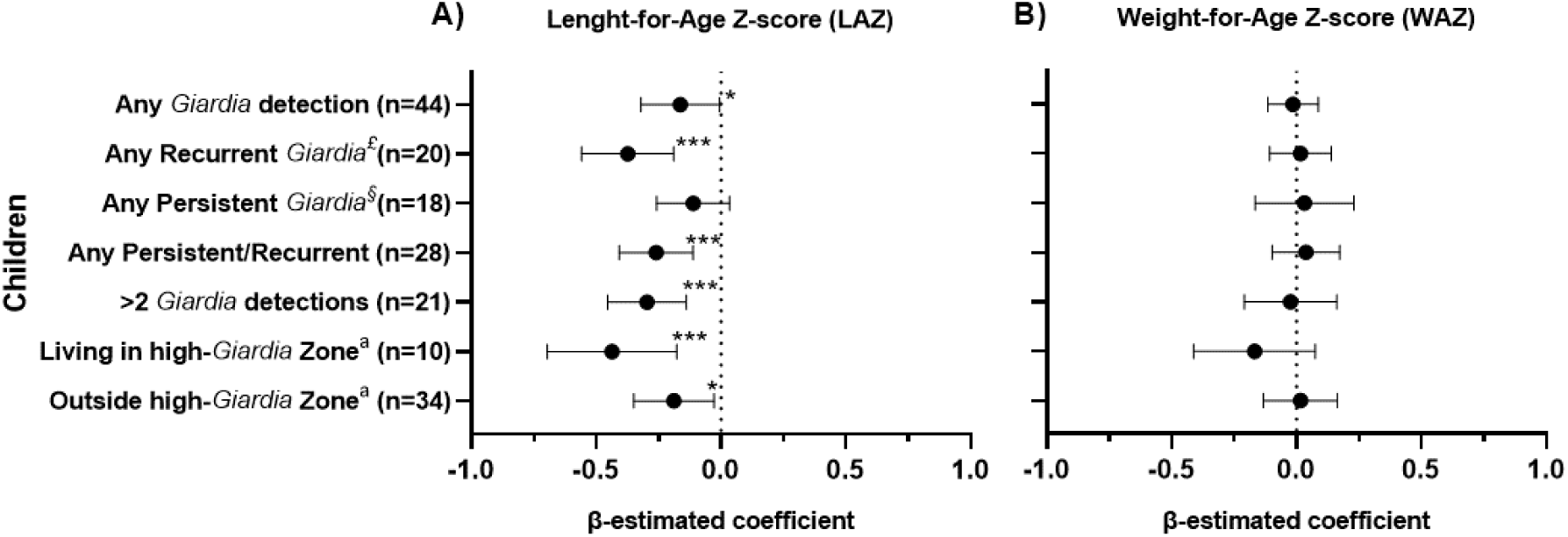
Estimation of linear regression in children infected with *Giardia* (n=44). A) Length-for-age (LAZ) and B) Weight-for-age (WAZ) Z-score in infected children. The β-estimated coefficient was calculated using generalized estimating equations (GEE), and the value one month prior to the *Giardia* infection was used as the baseline. Models were adjusted for age, mode of delivery, sex, socioeconomic status, breastfeeding, and episodes of diarrhea. ^£^Any recurrent infection was defined if a child had more than one *Giardia*-positive qPCR result in non-consecutive stool samples. ^§^Any persistent *Giardia* infections were defined if two or more consecutive routine stool samples that tested positive for *Giardia*. ^a^Children living within or outside high-*Giardia* burden areas based on the Kernel density distribution of *Giardia* infections. *P<0.050 to 0.010, **P<0.010 to 0.001, \*\*\**P*<0.001.

### *Giardia* does not associate with sustained increases in fecal biomarkers of EED

Fecal NEO, MPO, and Reg-1β concentrations were analyzed in the 58 children who had samples available at both 24 and 36 months of age (Fig. 4 and Table S4). All fecal biomarker concentrations were non-normally distributed at both time points. Total fecal protein was also quantified per sample and used to normalize biomarker concentrations (Fig. S6). Fecal biomarkers were compared between children with any prior *Giardia* detection (24/58 at 24 months and 31/58 at 36 months) and those without *Giardia* detection (Fig. 4A-4C). Only mean NEO at age 36 months was significantly lower (209.1 vs 335.4 nmol/L, *P*=0.024) in children with *Giardia* infections (Fig. 4B).

**Figure 4.**
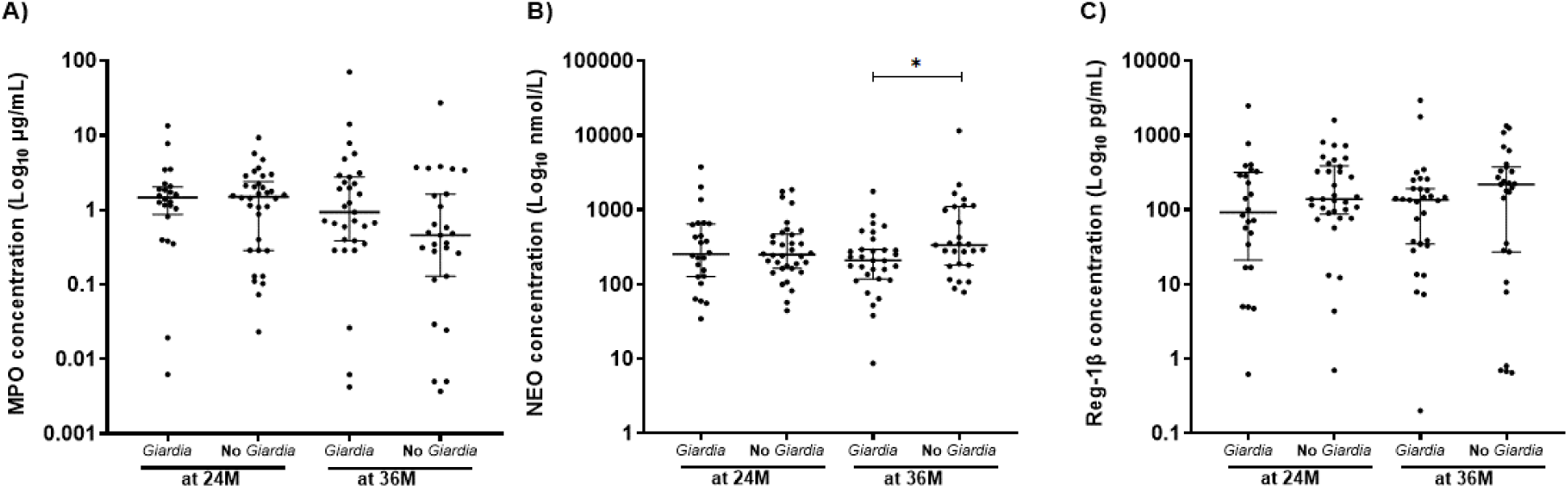
Fecal biomarkers measured in children with and without *Giardia* infections. Fecal biomarkers measured at 24 (24M) and 36 months of age (36M) in children infected at least once with *Giardia* infections at 24M (*Giardia*, n=24) and 36M (n=31), and children not infected at 24M (No *Giardia*, n=34) and 36M (n=27) for: A) Myeloperoxidase (MPO), B) Neopterin (NEO), C) Regenerating family member 1β (Reg-1β). \**P*<0.050 to 0.010, \*\**P*<0.010 to 0.001, \*\*\**P*<0.001.

### Association between *Giardia* infection and systemic biomarkers

Systemic biomarkers were evaluated at 24 months of age, and compared between children who had at least one *Giardia* detection regardless of symptoms (n=29) and those without any *Giardia* detection (n=20) (Fig. 5 and Table S5). The systemic biomarker profiles of children with *Giardia* detection during the first 24 months were characterized by higher concentrations of Anti-FliC IgA (9.8 [IQR: 7.9-11.2] vs 8.2 [IQR: 6.9-9.1] ng/L, *P*=0.033) (Fig. 5A), and I-FABP (7,074.8 [IQR: 4,271.4-11,085.8] vs 4,573 [IQR: 1,807.4-5,823.7] pg/mL, *P*=0.002) (Fig. 5B), and lower concentrations of IGF-1 (0.1 [IQR: 0.1-31.3] vs 203.3 [IQR: 0.09-433.6] ng/mL, *P*=0.005) (Fig. 5C), compared to children without *Giardia* detection. Both IGF-1 and I-FABP were associated with LAZ at 24 months of age. No significant associations were observed for CRP, RBP4, AGP, FGF21, sTfR, and sCD14 (Fig. 5D-5I).

**Figure 5.**
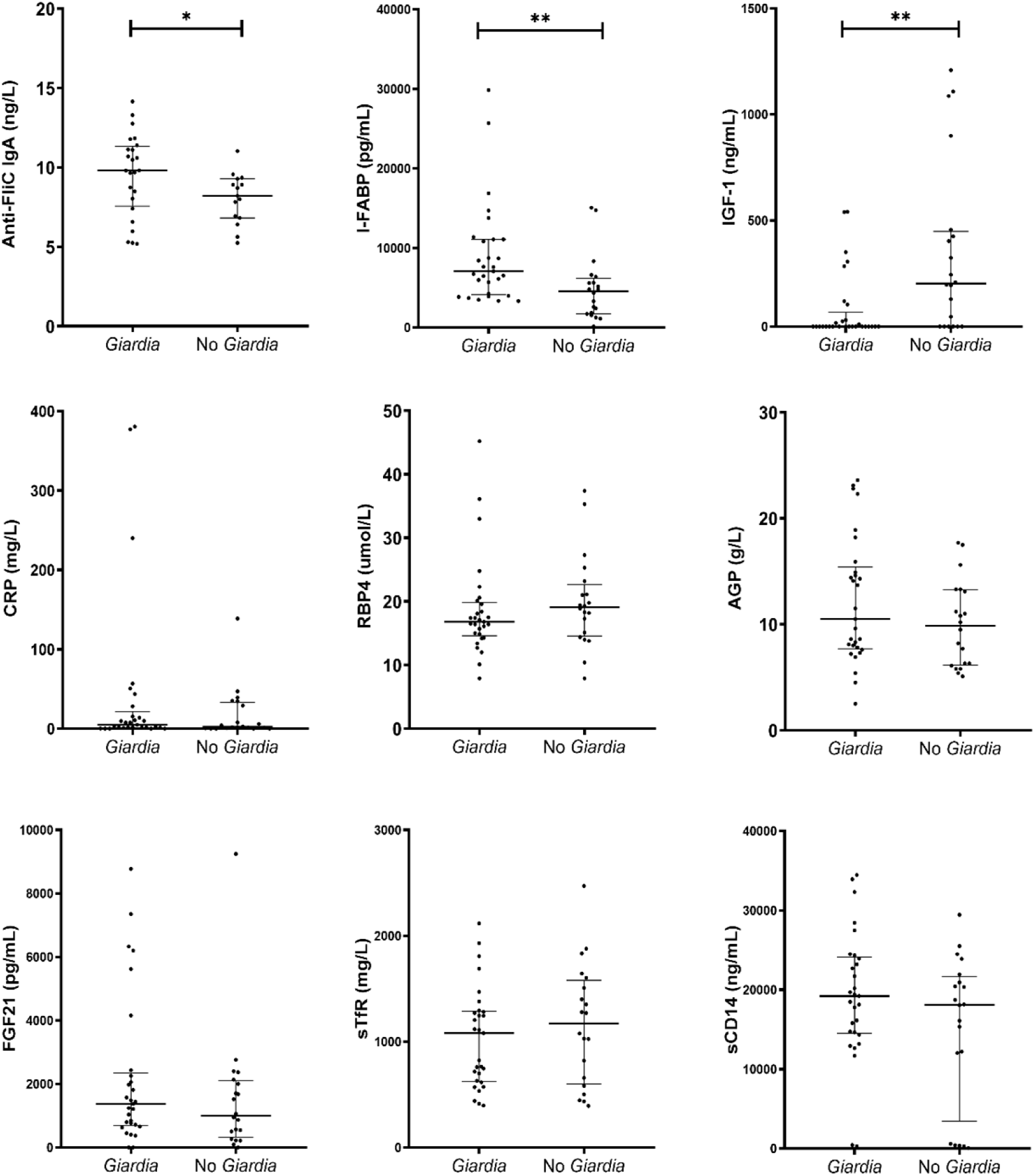
Systemic biomarkers measured at 24 months of age in children infected at least once with *Giardia* infections (*Giardia*, n=29) and children not infected (No *Giardia*, n=20). A) Anti-flagellin C (Anti-FliC)-IgA. B) Insulin-like growth factor-1 (IGF-1). C) Intestinal fatty acid binding protein (I-FABP). D) Fibroblast growth factor 21 (FGF21). E) Soluble transferrin receptor (sTfR). F) C-reactive protein (CRP). G) Retinol binding protein 4 (RBP4). H) α-1-acid glycoprotein (AGP). I) Soluble cluster of differentiation 14 (sCD14). \**P*<0.050 to 0.010, \*\**P*<0.010 to 0.001, \*\*\**P*<0.001.

In linear regression models, for every doubling of IGF-1, there was an average of 0.07 standard deviation increase in LAZ (95% CI: 0.005-0.136, p = 0.036). Conversely, for every doubling of AGP, an average of-0.694 standard deviation decrease in LAZ (95% CI:-1.250 to-0.134, p = 0.016) and every doubling of I-FABP, an average of-0.273 standard deviation decrease in LAZ (95% CI:-0.538 to-0.008, p = 0.044) (Table S6).

### Outcomes correlate with *Giardia* infection

We evaluated the correlation between the number of AGE episodes of any cause, AGE episodes with *Giardia* detection (GAGE), and the frequency of *Giardia* detection throughout the surveillance period, regardless of symptoms, with the anthropometric measurements, and biomarkers (Fig. 6). When analyzing the number of AGE episodes of any cause, no significant correlation was found between biomarkers and anthropometric measurements, except for WAZ at 3 years, which showed a weak negative correlation (rho:-0.226, P=0.049), indicating that increasing AGE by any cause, WAZ tends to decrease slightly. In contrast, when increasing number of GAGE events, a moderate positive corretation was observed for levels of I-FABP (rho: 0.462, *P*<0.001) and Anti-FliC IgA (rho: 0.311, *P*=0.043) levels, and weak negative correlation with LAZ at 2 years (rho:-0.236, *P*=0.040) (Fig. 6). Finally, correlations were also assessed using the total number of *Giardia* detections, regardless of symptoms, showing a moderate negative correlation with IGF-1 (rho:-0.433, *P*=0.002), NEO (rho:-0.274, *P*=0.037), and LAZ at 2 years (rho:-0.277, *P*=0.015), and positive correlations with I-FABP (rho: 0.506, *P*<0.001) and MPO (rho: 0.261, *P*=0.048).

**Figure 6.**
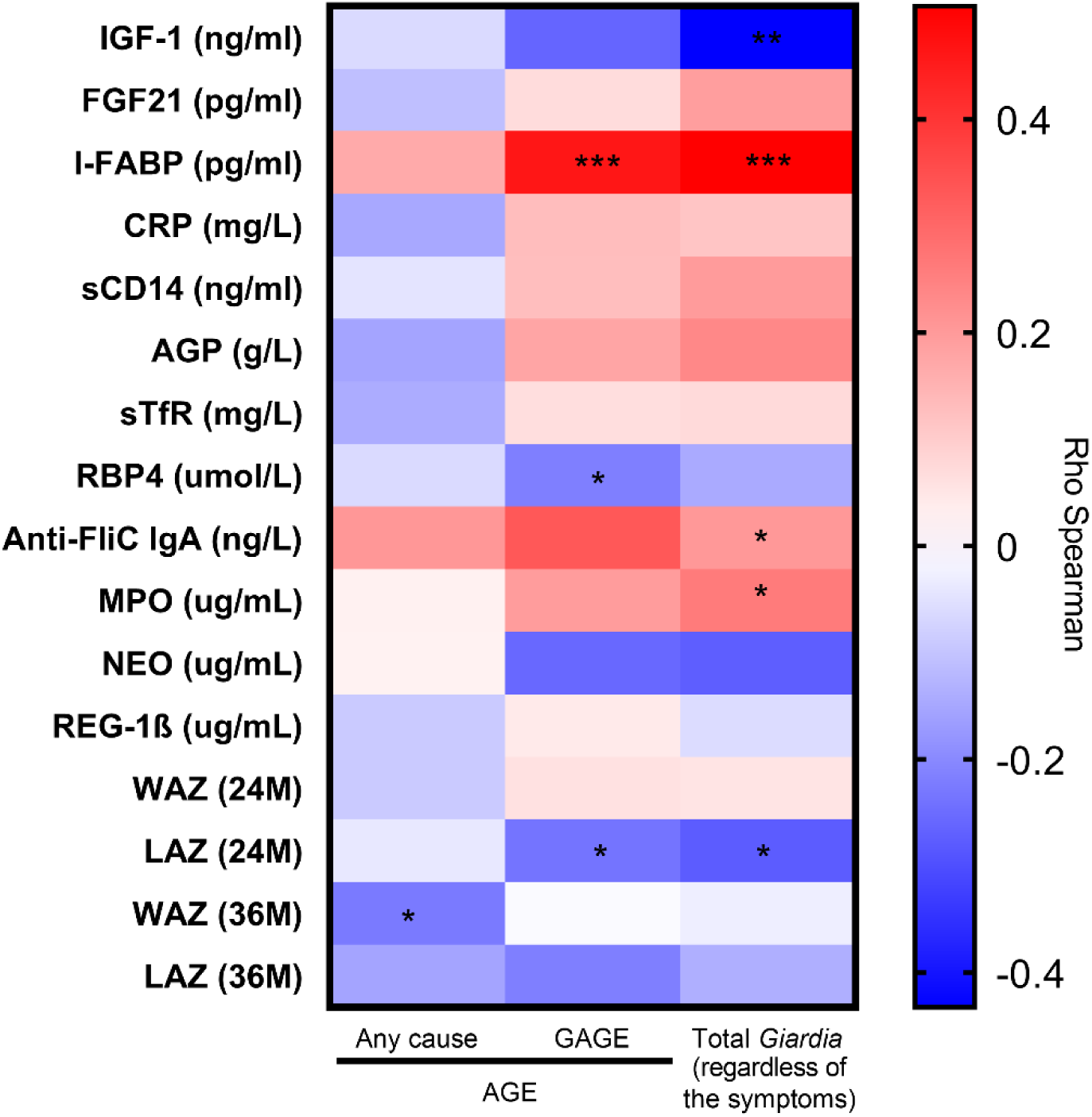
**Correlation between fecal and systemic biomarkers** with total acute gastroenteritis by any cause (Any cause), total of AGE episodes *Giardia-*positive (GAGE), and the total number of *Giardia* infections (regardless of symptoms). Correlations were performed using Spearman’s rank correlation with all episodes or infections reported in 49 children throughout the cohort for anti-flagellin C (Anti-FliC)-IgA, insulin-like growth factor-1 (IGF-1), intestinal fatty acid binding protein (I-FABP), fibroblast growth factor 21 (FGF21), soluble transferrin receptor (sTfR), C-reactive protein (CRP), retinol binding protein 4 (RBP4), α-1-acid glycoprotein (AGP), Length-for-age (LAZ), and Weight-for-age (WAZ) Z-score. *P<0.050 to 0.010, **P<0.010 to 0.001, ***P<0.001.

## Discussion

We leveraged a birth cohort study from León, Nicaragua [15] to study the natural history of *Giardia* infections and their impact on child health. This birth cohort provides data to study early-life *Giardia* infections, including epidemiology, growth measurements, and fecal and systemic biomarkers. We previously reported that *Giardia* was found in 7.5% of AGE episodes from the entire cohort data [16]. In this study, we focused our examination on the subcohort of children who provided complete monthly surveillance data (stool samples and demographics) to estimate the longitudinal effect of *Giardia* on growth outcomes and biomarkers of intestinal responses. By the end of the 36-month surveillance period, we found that 57.9% of children had at least one *Giardia* infection, with acquisitions beginning around 12 months of age, and incidence progressively increasing through the second year of life. Most of the first *Giardia* detections occurred in surveillance stools rather than in diarrheal stools, and persistent and repeated infections were common.

Consistent with previous reports linking *Giardia* to poverty and inadequate sanitation systems [23,24], our findings indicate that household-level deprivations, specifically the absence of basic needs and the presence of earthen floors, are strongly associated with an increased probability of *Giardia* infection, independent of seasonality, with this relationship being more pronounced among children with more than two infections. These results are similar to data from this cohort reported previously, where AGE with *Giardia* detection (GAGE) was associated with living in a household with a latrine and earthen floor [16], supporting that household environmental conditions may be an important determinant of the burden of exposure to the parasite. We detected *Giardia* during all months of the year, including during both rainy and dry seasons. Our findings suggest that *Giardia* infections were more prevalent in a specific geospatial area. We also identified a high-density area of *Giardia* infections characterized by low socioeconomic status and predominant use of latrines, which may contribute to the increased burden of *Giardia* in this area. Although further analysis could be conducted to explore the influence of different bandwidth settings on the density distribution, our results are in line with the finding that the geospatial distribution of the intestinal parasitic infections is influenced by socioeconomic conditions [24].

These findings highlight the relevance of geospatial and neighborhood contexts for both the frequency and incidence of infection, and point to potential opportunities for interventions that could reduce the burden of *Giardia* in this area and limit its spread to surrounding communities, as such community-level measures are likely to achieve larger and more durable reductions in transmission than isolated, household-level efforts alone. At the community level, these include health education, promotion of personal hygiene practices, and improvements in sanitation to reduce exposure to [25–27].

Our results show a significant *Giardia-*associated decrease of LAZ at 24 and 36 months of age, regardless of symptoms. There was a greater, nearly two-fold, impact in those children in whom *Giardia* was detected more than 2 times, including those with persistent or recurrent infections. Our findings are consistent with other studies in LMICs, which have found that *Giardia* is negatively associated with childhood linear growth [5,6,8–10]. Although we found that *Giardia* was associated with a decrease in LAZ. On the other hand, neopterin decreased significantly after *Giardia* infection. Similar findings were observed when comparing children who were infected with *Giardia* vs those without *Giardia*. Our study adds to emerging and consistent findings that *Giardia* does not produce intestinal inflammation during infection [4,13,14], despite associating with impaired linear growth. These findings suggest that impaired linear growth associated with *Giardia* infections appears to be a process independent of typical EED-like inflammation, and could be due to the direct disruptions in epithelial cells by parasite factors [28], including secreted proteins and the unique metabolic properties of the protozoan, and/or an interaction with resident intestinal microbes or other gut pathogens [12].

Interestingly, although there was no evidence of typical intestinal inflammation, systemic biomarkers like I-FABP and anti-FliC IgA were increased in children with at least one *Giardia* detection, regardless of the symptoms. These findings are consistent with separately reported associations of increased I-FABP [29] and anti-FliC IgA [6] levels in children following *Giardia* infection, supporting that *Giardia* is associated with intestinal epithelial disruption through mechanisms independent of conventional fecal EED biomarkers. On the other hand, IGF-1 was decreased in *Giardia*-infected children. To our knowledge, this is the first study to suggest that Giardia infection is associated with decreased systemic IGF-1 concentration. While IGF-1 is a well-recognized hormonal driver of linear growth, its relationship with *Giardia* infection has not been previously explored. This evidence suggests that *Giardia* may influence growth outcomes through pathways involving nutrient-metabolic disruption and endocrine modulation. A conceptual model suggests that malnutrition inhibits hepatic IGF-1 synthesis[30], this hypothesis was supported by mouse models in which IGF-1 levels are reduced during malnutrition [31].

Deprivation of amino acids has been also described as one of the different proposed mechanisms that could be associated with the effect on growth as part of the nutrient-metabolite disruption during *Giardia* infections [12]. Both essential and non-essential ammio acids have been reported to be decreased in some children infected with *Giardia* who concomitantly demonstrate poor growth [11]. This may arise through shortening of the intestinal brush-border microvilli, which could lead to lose the intestinal absorptive function [11,32–35], or altered amino acid availability due to parasite metabolism and/or interactions with intestinal microbiota. Amino acid deprivation has been shown to decrease IGF-1 mRNA expression in hepatocytes and muscle cells [36].

Additionally, certain amino acids, such as arginine, can specifically stimulate IGF-1 secretion and act through both growth hormone-dependent and independent pathways to regulate IGF-1 secretion [37].

This study has some limitations. First, the analysis was limited to a subset of 76 children from the SAGE cohort due resource restrictions. Second, during the third year of follow-up, stool sampling was performed quarterly rather than monthly, potentially reducing the sensitivity to detect incident infections. Third, detailed nutritional data were not collected in the cohort, including information on complementary feeding (food groups, and quantity), as well as food availability, access, consumption, and nutrient absorption. Additionally, the effects of specific pathogens were assessed only in the context of diarrheal episodes of any cause; specific pathogens were not included as covariates in the analysis. We also acknowledge the lack of adjustment for multiple comparisons (e.g., Benjamini–Hochberg test for false discovery rate), which may increase the likelihood of type I errors. Finally, systemic biomarker measurements were only available at the 2-year time point. Longitudinal data beyond two years would provide a better understanding of the durable impact of *Giardia* infection during early childhood.

Similarly, to obtain a comprehensive view of the endocrine axis, metabolic pathways, including growth hormone and insulin-like growth factor binding proteins, should be assessed.

In summary, our study provides an update of the natural history of *Giardia* infection in early childhood, to better understand timing of acquisition, association with symptoms, risk factors, geographical distribution, and its impact on child growth and EED. Our study also provided evidence that *Giardia* infection is negatively associated with LAZ, markers of chronic intestinal damage, and IGF-1 and positively associated with markers of intestinal epithelial disruption. The uncoupling of poor linear growth, epithelial disruption, and reduced growth signaling from intestinal inflammation seen in this study, other cohorts, and experimental models of giardiasis, is supporting a unique characteristic of endemic pediatric giardiasis that requires further investigation of its pathophysiology to guide future interventions.

## Data Availability

All relevant data are within the manuscript and its Supporting Information files

## Acknowledgments

Authors appreciate the support from all parents of the children participating in the SAGE cohort and recognize all efforts from the fieldwork team (Yorling Picado, Nancy Corea, Mileydis Soto, Maria Mendoza, Merling Balmaceda, Jhoseling Delgado, Ruth Neira, Veronica Pravia, Yuvielka Martinez, Aura Scott, Yadira Hernandez, Xiomara Obando (RIP) y Patricia Mendez) for collection of clinical and epidemiological information together with biological material. The support from the SILAIS-León in particular the personnel from the Perla Maria Norori Health Center. Additionally, we appreciate the support of the University of Costa Rica (UCR) under the project “Cooperation with the University of North Carolina for training and scientific dissemination in infectious diseases in Central America” project and the Doctoral Program in Sciences at UCR.

## Author Contributions

**Conceptualization:** Lester Gutiérrez, Luther A. Bartelt, Filemon Bucardo, Nadja A. Vielot, Sylvia Becker-Dreps, Samuel Vilchez

**Data curation:** Lester Gutiérrez, Nadja A. Vielot, Christian Toval-Ruiz, Roberto Herrera, Yaoska Reyes, Rebecca Barney.

**Formal analysis:** Lester Gutiérrez, Luther A. Bartelt, Nadja Vielot, Sylvia Becker-Dreps, Samuel Vilchez, Filemón Bucardo.

**Methodology:** Lester Gutiérrez, Nadja Vielot, Luther A. Bartelt, Filemon Bucardo, Sylvia Becker-Dreps, Samuel Vilchez

**Supervision:** Luther A. Bartelt, Nadja Vielot, Javier Mora, Sylvia Becker-Dreps, Samuel Vilchez, Robert KM Choy.

**Writing – original draft:** Lester Gutiérrez

**Writing – review & editing:** Luther A. Bartelt, Nadja Vielot, Javier Mora, Michael B. Arndt, Filemon Bucardo, Sylvia Becker-Dreps, Samuel Vilchez, Rebecca Barney, Robert KM Choy.

## Supporting information

**Table S1.** Baseline epidemiological characteristics of the sub-cohort versus the remaining cohort

**Table S2.** Biomarkers Associated with Growth, Systemic Inflammation, and Nutritional Status

**Table S3.** β-estimated coefficient of linear regression using GEE on child anthropometric indicators in children infected with *Giardia* (n=44).

**Table S4.** Fecal biomarkers measured at 24 and 36 months of age in children infected at least once with *Giardia* infections and children not infected.

**Table S5.** Systemic biomarkers measured at 24 months of age in children infected at least once with *Giardia* infections (*Giardia*, n=29) and children not infected (No *Giardia*, n=20).

**Table S6.** β-estimates from linear regression models of log2-transformed fecal and systemic biomarker concentrations on child anthropometric indicators at 24 months of age.

**Figure S1.** Timeline of routine stool collections, anthropometric assessment (weight-for-age [WAZ] and length-for-age [LAZ]), and timepoints of fecal and systemic biomarker measurements.

**Figure S2. Causal diagram used for adjusting for potential confounders.** Yellow circles with a triangular bullet (‣) represent exposure variables. The red circle represents an ancestor of exposure and outcome. Blue circles with a blue border represent an ancestor of the outcome. Blue circles with black borders represent the outcome. The green line means causal path, and the red line means bias pathway

**Figure S3.** Diagram illustrating the linear regression models using generalized estimating equations (GEE) to assess the effect of *Giardia* infection on Length-for-Age (LAZ) and Weight-for-Age (WAZ) Z score. Effects were estimated using monthly longitudinal LAZ and WAZ measurements. The model for any *Giardia* infection began at the first positive *Giardia* infection (red circle “b”), using the value one month prior as the baseline (green triangle “a”). The recurrent event models began at the second non-consecutive *Giardia* positive stool (red circle “c”), with the baseline defined as one month prior to this event (green triangle). The persistent event model started at the second consecutive positive stool (red circle “d”), with the baseline defined as one month prior to this event (green triangle). All models were adjusted for child’s age, mode of delivery, sex, socioeconomic status, breastfeeding, and episodes of diarrhea during the same period. *A *P* value <0.05 was considered statistically significant

**Figure S4. Timing of *Giardia* infections in surveillance and diarrheal stools among 76 children.** The Y-axis represents child ID, while the X-axis represents the child’s age in months. *Giardia* detection in stools samples was determined using qPCR. Light gray squares: samples with no *Giardia* detection. Yellow squares: surveillance stool samples with a positive *Giardia* detection. Red squares: diarrheal stool samples with positive *Giardia* detection. White squares: monthly stools samples that were not collected. Dark gray squares: diarrheal stools with a *Giardia* negative detection.

**Figure S5. Distribution of *Giardia* infections in 76 children during the first 3 years of life.** A) Monthly absolute distribution from June 2017 to March 2021. B) Frequency of *Giardia* positive stools by season, rainy (from March to September) and dry season (from November to April). C) Spatial distribution of 76 children included in the study, red points represent children infected with *Giardia* (n=44). D) Kernel Density Distribution of *Giardia* infections. Darker area represents geographic zone (km^2^) with higher concentrations of infection events.

**Figure S6. Fecal biomarkers measured at 24 and 36 months of age** in children infected at least once with Giardia infections at 24 (Giardia, n=24) and 36 months of age old (n=31), and children not infected at 24M (No Giardia, n=34) and 36M (n=27) normalized to total fecal proteins (A) for: B) Neopterin (NEO). C) Myeloperoxidase (MPO), and D) Regenerating family member 1β (Reg-1β). *P<0.050 to 0.010, **P<0.010 to 0.001, ***P<0.001.

## File naming supporting information

Supporting information was saved as “Supplemental_Doc.docx”

